# The association between plasma metabolites and sleep quality in the Southall and Brent Revisited Study (SABRE): A cross-sectional analysis

**DOI:** 10.1101/2020.07.30.20165217

**Authors:** Constantin-Cristian Topriceanu, Therese Tillin, Nishi Chaturvedi, Roshni Joshi, Victoria Garfield

## Abstract

**Background:** Disordered metabolic processes have been associated with abnormal sleep patterns. However, the biological triggers and pathways are yet to be elucidated.

**Methods:** Participants were from the Southall and Brent REvisited (SABRE) cohort. Nuclear Magnetic Resonance spectroscopy provided 146 circulating plasma metabolites. Sleep questionnaires identified the presence or absence of: difficulty falling asleep (DFA), early morning waking (EMW), waking up tired (WUT) and snoring. Metabolites were compared between the sleep quality categories using the t-test, then filtered using a false discovery rate of 0.05. Generalized linear models with logit-link assessed the associations between filtered metabolites and sleep phenotypes. Adjustment was made for important demographic and health-related covariates.

**Results:** 2718 SABRE participants were included. After correcting for multiple testing, 3 metabolites remained for DFA, 59 for snoring and none for EMW and WUT. In fully-adjusted models, 1 standard deviation increase in serum histidine, leucine and valine associated with lower odds of DFA by 0.84-0.89 (95% confidence intervals [CIs]: 0.75-0.99). Branched chain amino acids (ORs 1.11-1.15, 95%CIs 1.01-1.26) were positively associated with snoring. Docosahexaenoic acid (DHA) (OR 0.89, 95% CI 0.82-0.96) and total cholesterol in low-density lipoprotein (LDL) (OR 0.89, 95% CI 0.82-0.96) and high-density lipoprotein (HDL) (ORs 0.90, 95% CIs 0.83-0.99) associated with lower odds of snoring.

**Conclusion:** Histidine, leucine and valine associated with lower odds of difficulty falling asleep, while docosahexaenoic acid and cholesterol LDL and HDL subfractions associated with lower odds of snoring. Identified metabolites could provide guidance on the metabolic pathways behind the adverse sleep quality.

**Key messages:**

- Higher histidine, leucine and valine associate with lower odds of difficulty falling asleep.
- Branched amino acids (isoleucine, leucine and valine) were higher in participants experiencing snoring.
- Total cholesterol in LDL and HDL appeared to be beneficial in terms of snoring.

## 1. INTRODUCTION

Sleep is a vital component of the human circadian rhythm. Abnormal sleep patterns are increasingly common. Two of the most prevalent sleep disorders are insomnia and snoring, with recent estimates suggesting insomnia is reported in more than 30% and, snoring in more than 20% of the global population ^1^.

Sleep is of primordial importance in physiological homeostasis. While adverse sleep phenotypes are associated with negative health consequences, including cardiovascular disease and cancer ^2^, the triggers and pathways that may contribute to abnormal sleep phenotypes and related detrimental health outcomes have yet to be elucidated.

Metabolic dysfunction has been previously associated with sleep phenotypes ^3^. Insomnia and short sleep duration have been associated with increased odds of developing type 2 diabetes^4^, which could be mediated by branched chain amino acids^5^. Snoring has been associated with disordered metabolic processes including insulin resistance, hyperglycaemia and dyslipidemia (high triglycerides, high LDL cholesterol, low HDL cholesterol) ^6,7^. However, the directionality of the association between metabolites and sleep phenotypes is still unclear, but it appears that disordered sleep may be both a cause and a consequence of abnormal metabolism.

Thus, using a metabolomics approach^8^ could lead to a better understanding of the mechanisms underlying abnormal sleep phenotypes as well as provide a direction for future novel guided therapies. Similar approaches have been successfully employed, for example in characterizing novel predictors for heart failure^9^.

In this cross-sectional study, Nuclear Magnetic Resonance (NMR) spectroscopy was used to identify metabolites that are associated with sleep phenotypes; difficulty falling asleep, early morning waking, waking up tired and snoring reported in participants from the Southall and Brent REvisited (SABRE) cohort.

## 2. METHODS

### Participants and Study Design

The SABRE study is a tri-ethnic cohort of European, South Asian and African Caribbean participants living in West and North London (Southall and Brent districts). Between 1988-1991, participants aged between 40 to 69 years were randomly selected from 5-year age and sex stratified primary care lists (n=4.063) and workplaces (n=795). The full cohort details have been published elsewhere^5^. Ethnicity was self-assigned and agreed with the interviewer. South Asians and African Caribbeans were all first-generation migrants.

At baseline, participants were invited to a clinic appointment which involved completing a health and lifestyle questionnaire which included questions on sleep patterns. Fasting bloods were collected, and anthropometrics and blood pressure were measured. Diabetes was identified from self-report of physician diagnosis or receipt of anti-diabetes medications. In addition, oral glucose tolerance testing was performed. Undiagnosed diabetes was identified retrospectively using WHO 1999 criteria^10^.

The SABRE baseline study was granted ethics approval from Ealing, Hounslow and Spelthorne, Parkside and University College London research ethics committees.

### Exposures

Exposures were NMR measured 146 circulating plasma metabolites including amino acids, small molecules (e.g. glycerol, pyruvate) and a detailed lipid profile consisting of 16 lipoprotein subclasses together with their lipid (phospholipids, triglycerides, total and free cholesterol, and cholesterol esters) component concentrations and particle dimensions.

Serum fasting samples were obtained from participants at baseline and for the Southall centre only (n=3700) were stored at -80°C. In 2012, a proton NMR spectrum was employed to detect circulating plasma metabolite levels following the signal suppression of other molecules according to methodologies previously used ^11^. Ratios were excluded.

None of the study participants were receiving lipid-lowering medication at the time of the baseline studies.

### Outcomes

The outcomes of interest were four sleep quality phenotypes: difficulty falling asleep, early morning waking, waking up tired and snoring.

Participants were asked to rate their sleep quality in the past 30 days at visit 1 (1988-90) using four questions adapted from the validated Jenkins Sleep Questionnaire (JSEQ)^12^. They were asked whether they felt they had been waking up too early in the morning, had difficulties falling asleep at night, had problems with snoring during the night and whether they woke up feeling tired. All sleep phenotypes were rated binary (i.e. ‘No’/0 or ‘Yes’/1).

### Covariates

Covariates were selected a priori based on prior associations with sleep quality and metabolism. Covariates were recorded at the time of the baseline studies (when serum samples were collected for storage) and included: age, sex, ethnicity, years of education, waist-hip-ratio (WHR), cardiovascular disease (CVD; coronary artery disease and stroke), Type 2 diabetes (T2DM), hypertension medication, total number of alcohol units per week and smoking status (never smoked, ex-smoker, current smoker). CVD, T2DM and hypertension medication were recorded as binary (yes/no).

### Statistical methods

Data distribution was assessed graphically using histograms and statistically using the Shapiro-Wilk test. Continuous variables were expressed as medians (interquartile ranges), while categorical variables were expressed as counts (percentage). Metabolite concentrations were log-transformed, mean centered and scaled to a standard deviation (SD) of 1 before further analysis.

T-test analysis was used as a screening tool to identify metabolites linked with sleep phenotypes, correcting for multiple testing at a false discovery threshold of 0.05 ^13^. Metabolites that passed this threshold were further referred to as candidate metabolites. Candidate metabolite associations with sleep phenotypes were evaluated using multivariable generalized linear models accounting for age, sex, ethnicity and years of education (Model 1). Further adjustments were model 1 plus WHR, CVD, T2DM, anti-hypertensive medication, alcohol units and smoking status (model 2).

About a fifth of all participants reported zero weekly alcohol intake. To account for this high degree of zero-inflation, ^14^, a modified version of GLM (using template model builder [glmmTMB]) was chosen (**Supplementary Note 1**).Thus, glmmTMB with binomial distribution and logit link (equivalent to logistic regression) using metabolites as exposures was employed to predict the binary sleep phenotypes as outcomes.

The T-test is not able to discriminate between groups where minor differences exists. This may not identify biologically relevant metabolites^8^. Thus, we ran the regression models for all the available metabolites as a sensitivity analysis.

Statistical analysis was performed in R (version-3.6.0)^15^. P-values <0.05 were considered significant.

## 3. RESULTS

### PARTICIPANT CHARACTERISTICS

Of the 4,858 SABRE participants, 3,700 (from the Southall Centre only) had stored blood samples with 3,255 being viable for NMR analysis. Furthermore, 2,718 had all sleep phenotypes, covariates and metabolites. Of the 2,718 included participants, 23.79 (87.53%) were male and 664 (24.43%) were current smokers. The mean age of the cohort was 52 ± 7.18 and the mean WHR 0.95 ± 0.08. Difficulty falling asleep was reported in 453 (16.67%) participants, early morning waking in 1,108 (40.77%), waking up tired in 926 (34.11%) and snoring in 1051 (38.67%). Participants characteristics are summarized in **Table 1**.

**Table 1.** Baseline demographic characteristics of Southall And Brent Revisited (SABRE) cohort participants stratified by sleep phenotype status

| Sleep phenotype | Difficulty Falling Asleep |  |  | Early Morning Waking |  |  | Waking Up Tired |  |  | Snoring |  |  | Overall |
| --- | --- | --- | --- | --- | --- | --- | --- | --- | --- | --- | --- | --- | --- |
|  | Yes<br>453<br>(16.67) | No<br>2265<br>(83.33) | p-value | Yes<br>1108<br>(40.77) | No<br>1610<br>(59.23) | p-value | Yes<br>927<br>(34.11) | No<br>1791<br>(65.89) | p-value | Yes<br>1051<br>(38.67) | No<br>1667<br>(61.33) | p-value | n=2718 |
| Age, years | 52.08 ±<br>7.27 | 52.10 ±<br>7.16 | 0.966 | 52.15 ±<br>7.11 | 52.06 ±<br>7.23 | 0.709 | 51.25 ±<br>7.05 | 52.53 ±<br>7.20 | <0.0001 | 52.66 ±<br>7.01 | 51.74 ±<br>7.25 | 0.001 | 52.10 ±<br>7.18 |
| Male sex (%) | 361<br>(79.69) | 2018<br>(89.09) | <0.0001 | 961<br>(86.73) | 1418<br>(88.07) | 0.327 | 787<br>(84.89) | 1592<br>(89.06) | 0.004 | 982<br>(93.43) | 1397<br>(83.80) | <0.0001 | 2379<br>(87.53) |
| Ethnicity |  |  |  |  |  |  |  |  |  |  |  |  |  |
| European | 193<br>(42.60) | 1159<br>(51.17) | 0.002 | 468<br>(42.24) | 884<br>(54.91) | <0.0001 | 456<br>(49.19) | 896<br>(50.03) | 0.528 | 512<br>(49.57) | 840<br>(50.39) | 0.681 | 1352<br>(49.74) |
| South Asian | 238<br>(52.5) | 984<br>(43.44) |  | 591<br>(53.34) | 631<br>(39.19) |  | 427<br>(46.06) | 795<br>(44.39) |  | 481<br>(45.77) | 741<br>(44.45) |  | 1222<br>(44.96) |
| African-Caribbean | 22<br>(0.05) | 122<br>(5.39) |  | 49<br>(4.42) | 95<br>(5.9) |  | 44<br>(4.75) | 100<br>(5.58) |  | 58<br>(4.66) | 86<br>(5.16) |  | 144<br>(5.30) |
| Years of education | 10.97 ±<br>3.25 | 11.25 ±<br>3.11 | 0.083 | 11.11 ±<br>3.18 | 11.27 ±<br>3.11 | 0.197 | 11.14 ±<br>3.27 | 11.24 ±<br>3.07 | 0.461 | 11.07 ±<br>3.19 | 11.29 ±<br>3.11 | 0.062 | 11.21 ±<br>3.14 |
| Waist-hip-ratio | 0.94 ±<br>0.09 | 0.94<br>±0.08 | 0.864 | 0.95 ±<br>0.08 | 0.94 ±<br>0.08 | 0.0004 | 0.94 ±<br>0.09 | 0.95 ±<br>0.08 | 0.546 | 0.96 ±<br>0.08 | 0.93 ±<br>0.09 | <0.0001 | 0.95 ±<br>0.08 |
| Cardiovascular disease | 59<br>(13.02) | 184<br>(8.12) | 0.001 | 119<br>(10.74) | 124<br>(7.70) | 0.008 | 112<br>(12.08) | 131<br>(7.31) | <0.0001 | 114<br>(10.85) | 129<br>(7.74) | 0.007 | 243<br>(8.94) |
| Diabetes | 72<br>(15.89) | 276<br>(12.19) | 0.038 | 151<br>(13.63) | 187<br>(11.62) | 0.313 | 120<br>(12.95) | 228<br>(12.73) | 0.922 | 152<br>(14.46) | 196<br>(11.76) | 0.046 | 348<br>(12.80) |
| Antihypertensives | 80<br>(17.66) | 279<br>(12.32) | 0.003 | 171<br>(15.43) | 188<br>(11.68) | 0.005 | 149<br>(16.07) | 210<br>(11.72) | 0.002 | 165<br>(15.70) | 194<br>(11.63) | 0.003 | 359<br>(13.21) |
| **Alcohol consumption, units** | 12.14 ± 17.63 | 11.67 ± 15.24 | 0.601 | 11.94 ± 15.56 | 11.62 ± 15.01 | 0.610 | 12.15 ± 16.58 | 11.54 ± 15.16 | 0.351 | 12.71 ± 16.78 | 11.14 ± 14.88 | **0.013** | 11.75 ± 15.66 |
| **Smoking status** |  |  |  |  |  |  |  |  |  |  |  |  |  |
| **Never** | 231<br>(50.99) | 1211<br>(53.47) | **0.008** | 622<br>(56.14) | 820<br>(50.93) | **0.027** | 482<br>(52.00) | 960<br>(48.71) | 0.066 | 518<br>(49.29) | 924<br>(55.43) | **0.006** | 1442<br>(53.05) |
| **Ex-smoker** | 96<br>(21.19) | 568<br>(25.08) |  | 231<br>(20.85) | 381<br>(23.66) |  | 232<br>(25.00) | 380<br>(21.22) |  | 250<br>(23.79) | 362<br>(21.72) |  | 612<br>(22.52) |
| **Current smoker** | 126<br>(27.82) | 486<br>(21.45) |  | 255<br>(23.01) | 409<br>(25.41) |  | 213<br>(23.00) | 451<br>(25.18) |  | 283<br>(26.92) | 381<br>(22.85) |  | 664<br>(24.43) |

Characteristics of participants stratified by sleep phenotypes. Values are presented as mean ± standard deviation for continuous variables or n (%) for categorical variables. P-values were calculated using t-test for continuous variables or $\chi^2$ -test for categorical variables.

### SCREENING FOR CANDIDATE METABOLITES

Of the 146 available metabolites, 12 candidate metabolites were identified for difficulty falling asleep, 4 for early morning waking, 2 for waking up tired and 73 for snoring (**Supplementary Table S1**). After correction for multiple testing, histidine, leucine and valine remained for difficulty falling asleep. 59 metabolites were further considered as candidates in association with snoring. None remained for early morning waking and waking up tired (**Supplementary Table S1)**.

### DIFFICULTY FALLING ASLEEP

After adjusting for model 1, serum histidine, leucine and valine were inversely associated with difficulty falling asleep. In the fully-adjusted models, 1 SD increases in serum histidine, leucine and valine were associated with lower odds of difficulty falling asleep by odds ratio (OR) 0.89 (95% confidence interval [CI]: 0.80 to 0.99), 0.84 (95% CI 0.75 to 0.96), and 0.84 (0.75 to 0.94), respectively.

### SNORING

Of 59 candidate metabolites associated with snoring, 31 remained associated with snoring in the fully-adjusted models (**Figure 1**). The odds ratios associated with 1SD increments in each of the 31 metabolites in Model 2 are presented in **Table 2** and visually illustrated in **Figure 2**. Amino acids and small molecules (e.g. acetate) were associated with higher odds of snoring. The largest positive estimate was observed for the aromatic amino acid, phenylalanine: OR 1.16 (95% CI 1.06-1.26). All branched-chain amino acids (BCCA) (isoleucine, leucine and valine) were associated with greater odds of snoring (OR in the range: 1.11-1.15). Serum lipids and lipoproteins (e.g. apolipoprotein (apo) A-I, docosahexaenoic acid, polyunsaturated fatty acids) (OR 0.88-0.92, 95% CIs 0.81-0.99) and total cholesterol in high-density lipoprotein (HDL) (OR 0.90, 95% CI 0.83-0.99) and in low-density lipoprotein (LDL) (OR 0.89, 95% CI 0.82-0.96) appeared to be beneficial. Regarding the subfraction breakdown, total cholesterol in small HDL (OR 0.91, 95% CI 0.84-0.99) and in small, medium and large LDL (ORs 0.87-0.90, 95% CI 0.80-0.97) associated with lower odds of snoring. In addition, cholesterol-esters and phospholipids from certain LDL and HDL subfractions were also inversely associated with snoring (**Figure 2**), with the largest OR for lower odds of snoring being observed for cholesterol-esters in medium-LDL (OR 0.64, 95% CI 0.45-0.92).

**Table 2.**
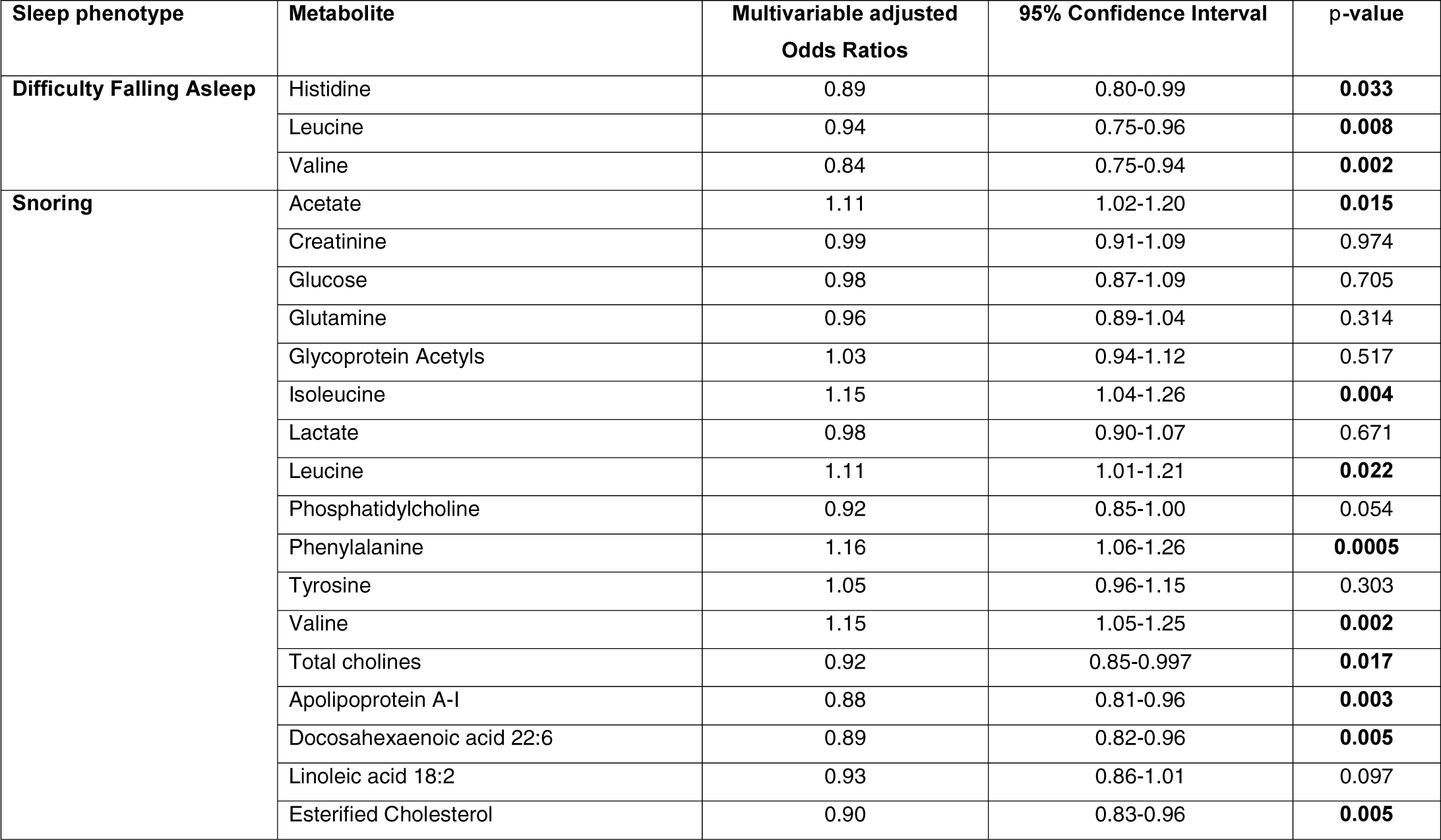
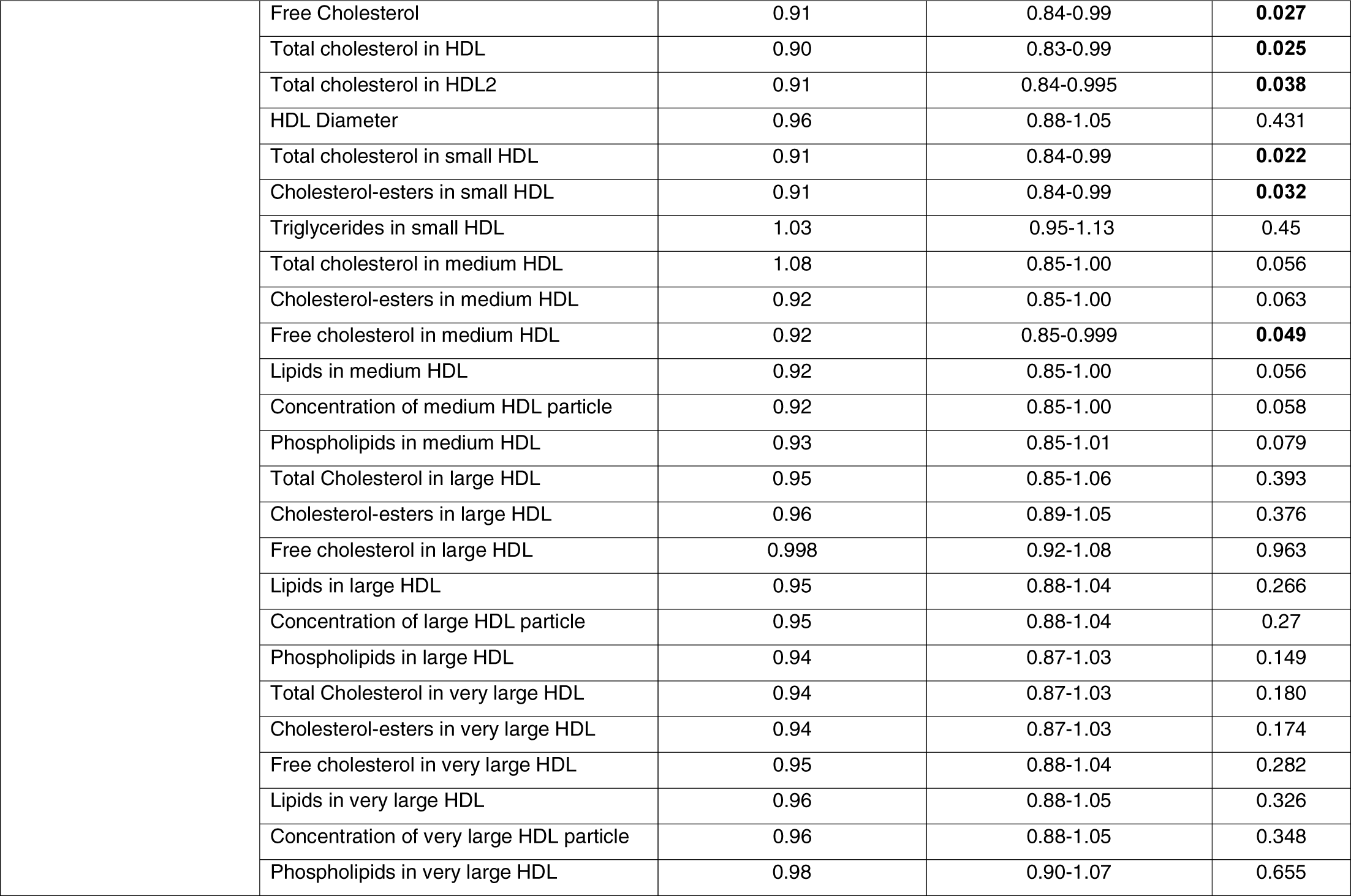
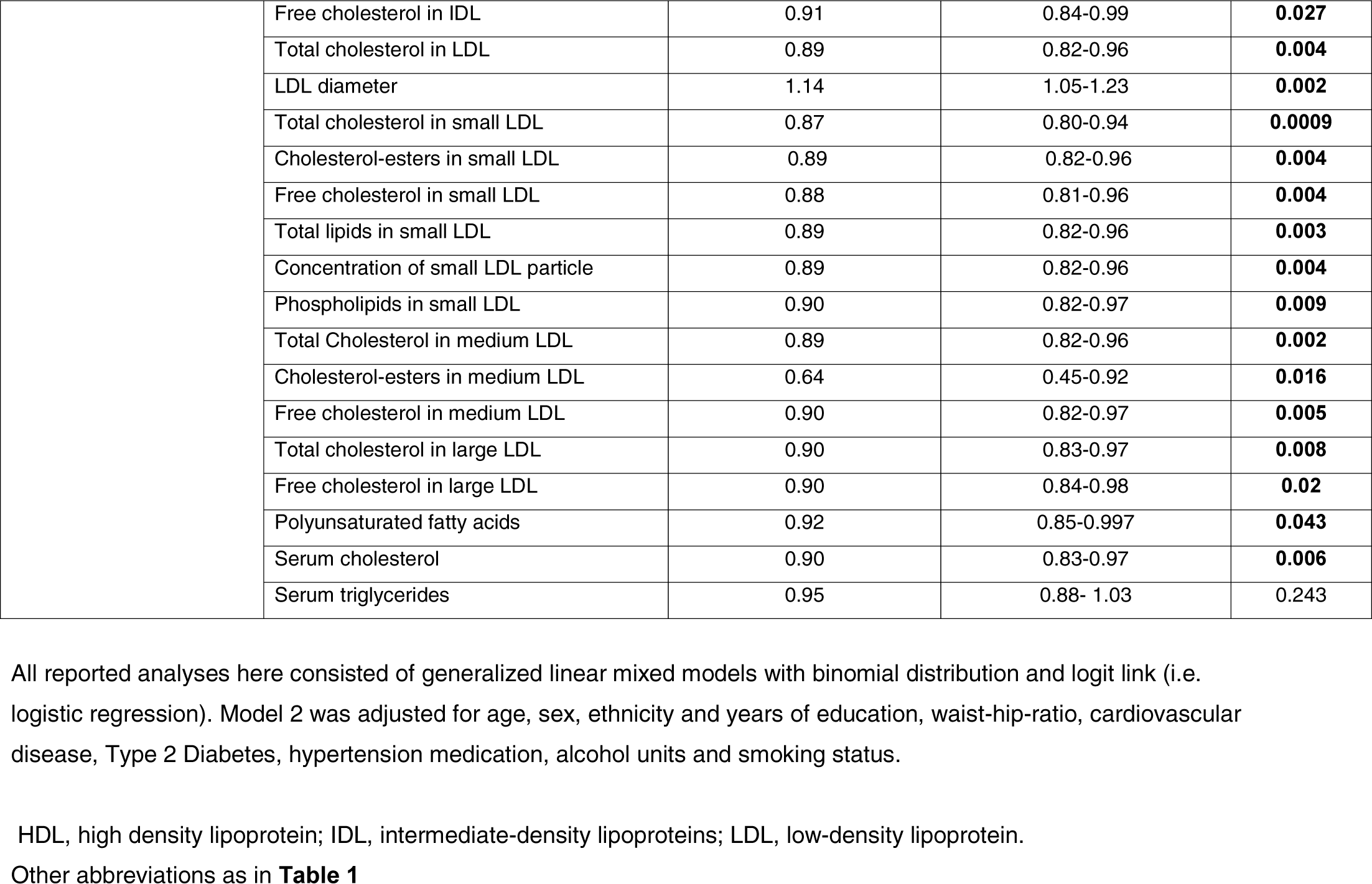
Associations between the metabolites who have passed the screening stage and the sleep phenotypes.

| **Sleep phenotype** | **Metabolite** | **Multivariable adjusted**<br>**Odds Ratios** | **95% Confidence Interval** | **p-value** |
| --- | --- | --- | --- | --- |
| **Difficulty Falling Asleep** | Histidine | 0.89 | 0.80-0.99 | **0.033** |
|  | Leucine | 0.94 | 0.75-0.96 | **0.008** |
|  | Valine | 0.84 | 0.75-0.94 | **0.002** |
| **Snoring** | Acetate | 1.11 | 1.02-1.20 | **0.015** |
|  | Creatinine | 0.99 | 0.91-1.09 | 0.974 |
|  | Glucose | 0.98 | 0.87-1.09 | 0.705 |
|  | Glutamine | 0.96 | 0.89-1.04 | 0.314 |
|  | Glycoprotein Acetyls | 1.03 | 0.94-1.12 | 0.517 |
|  | Isoleucine | 1.15 | 1.04-1.26 | **0.004** |
|  | Lactate | 0.98 | 0.90-1.07 | 0.671 |
|  | Leucine | 1.11 | 1.01-1.21 | **0.022** |
|  | Phosphatidylcholine | 0.92 | 0.85-1.00 | 0.054 |
|  | Phenylalanine | 1.16 | 1.06-1.26 | **0.0005** |
|  | Tyrosine | 1.05 | 0.96-1.15 | 0.303 |
|  | Valine | 1.15 | 1.05-1.25 | **0.002** |
|  | Total cholines | 0.92 | 0.85-0.997 | **0.017** |
|  | Apolipoprotein A-I | 0.88 | 0.81-0.96 | **0.003** |
|  | Docosahexaenoic acid 22:6 | 0.89 | 0.82-0.96 | **0.005** |
|  | Linoleic acid 18:2 | 0.93 | 0.86-1.01 | 0.097 |
|  | Esterified Cholesterol | 0.90 | 0.83-0.96 | **0.005** |

|  |  |  |  |
| --- | --- | --- | --- |
| Free Cholesterol | 0.91 | 0.84-0.99 | **0.027** |
| Total cholesterol in HDL | 0.90 | 0.83-0.99 | **0.025** |
| Total cholesterol in HDL2 | 0.91 | 0.84-0.995 | **0.038** |
| HDL Diameter | 0.96 | 0.88-1.05 | 0.431 |
| Total cholesterol in small HDL | 0.91 | 0.84-0.99 | **0.022** |
| Cholesterol-esters in small HDL | 0.91 | 0.84-0.99 | **0.032** |
| Triglycerides in small HDL | 1.03 | 0.95-1.13 | 0.45 |
| Total cholesterol in medium HDL | 1.08 | 0.85-1.00 | 0.056 |
| Cholesterol-esters in medium HDL | 0.92 | 0.85-1.00 | 0.063 |
| Free cholesterol in medium HDL | 0.92 | 0.85-0.999 | **0.049** |
| Lipids in medium HDL | 0.92 | 0.85-1.00 | 0.056 |
| Concentration of medium HDL particle | 0.92 | 0.85-1.00 | 0.058 |
| Phospholipids in medium HDL | 0.93 | 0.85-1.01 | 0.079 |
| Total Cholesterol in large HDL | 0.95 | 0.85-1.06 | 0.393 |
| Cholesterol-esters in large HDL | 0.96 | 0.89-1.05 | 0.376 |
| Free cholesterol in large HDL | 0.998 | 0.92-1.08 | 0.963 |
| Lipids in large HDL | 0.95 | 0.88-1.04 | 0.266 |
| Concentration of large HDL particle | 0.95 | 0.88-1.04 | 0.27 |
| Phospholipids in large HDL | 0.94 | 0.87-1.03 | 0.149 |
| Total Cholesterol in very large HDL | 0.94 | 0.87-1.03 | 0.180 |
| Cholesterol-esters in very large HDL | 0.94 | 0.87-1.03 | 0.174 |
| Free cholesterol in very large HDL | 0.95 | 0.88-1.04 | 0.282 |
| Lipids in very large HDL | 0.96 | 0.88-1.05 | 0.326 |
| Concentration of very large HDL particle | 0.96 | 0.88-1.05 | 0.348 |
| Phospholipids in very large HDL | 0.98 | 0.90-1.07 | 0.655 |
|  | Free cholesterol in IDL | 0.91 | 0.84-0.99 | **0.027** |
|  | Total cholesterol in LDL | 0.89 | 0.82-0.96 | **0.004** |
|  | LDL diameter | 1.14 | 1.05-1.23 | **0.002** |
|  | Total cholesterol in small LDL | 0.87 | 0.80-0.94 | **0.0009** |
|  | Cholesterol-esters in small LDL | 0.89 | 0.82-0.96 | **0.004** |
|  | Free cholesterol in small LDL | 0.88 | 0.81-0.96 | **0.004** |
|  | Total lipids in small LDL | 0.89 | 0.82-0.96 | **0.003** |
|  | Concentration of small LDL particle | 0.89 | 0.82-0.96 | **0.004** |
|  | Phospholipids in small LDL | 0.90 | 0.82-0.97 | **0.009** |
|  | Total Cholesterol in medium LDL | 0.89 | 0.82-0.96 | **0.002** |
|  | Cholesterol-esters in medium LDL | 0.64 | 0.45-0.92 | **0.016** |
|  | Free cholesterol in medium LDL | 0.90 | 0.82-0.97 | **0.005** |
|  | Total cholesterol in large LDL | 0.90 | 0.83-0.97 | **0.008** |
|  | Free cholesterol in large LDL | 0.90 | 0.84-0.98 | **0.02** |
|  | Polyunsaturated fatty acids | 0.92 | 0.85-0.997 | **0.043** |
|  | Serum cholesterol | 0.90 | 0.83-0.97 | **0.006** |
|  | Serum triglycerides | 0.95 | 0.88- 1.03 | 0.243 |

All reported analyses here consisted of generalized linear mixed models with binomial distribution and logit link (i.e. logistic regression). Model 2 was adjusted for age, sex, ethnicity and years of education, waist-hip-ratio, cardiovascular disease, Type 2 Diabetes, hypertension medication, alcohol units and smoking status.
HDL, high density lipoprotein; IDL, intermediate-density lipoproteins; LDL, low-density lipoprotein.
Other abbreviations as in **Table 1**

**Figure 1.**
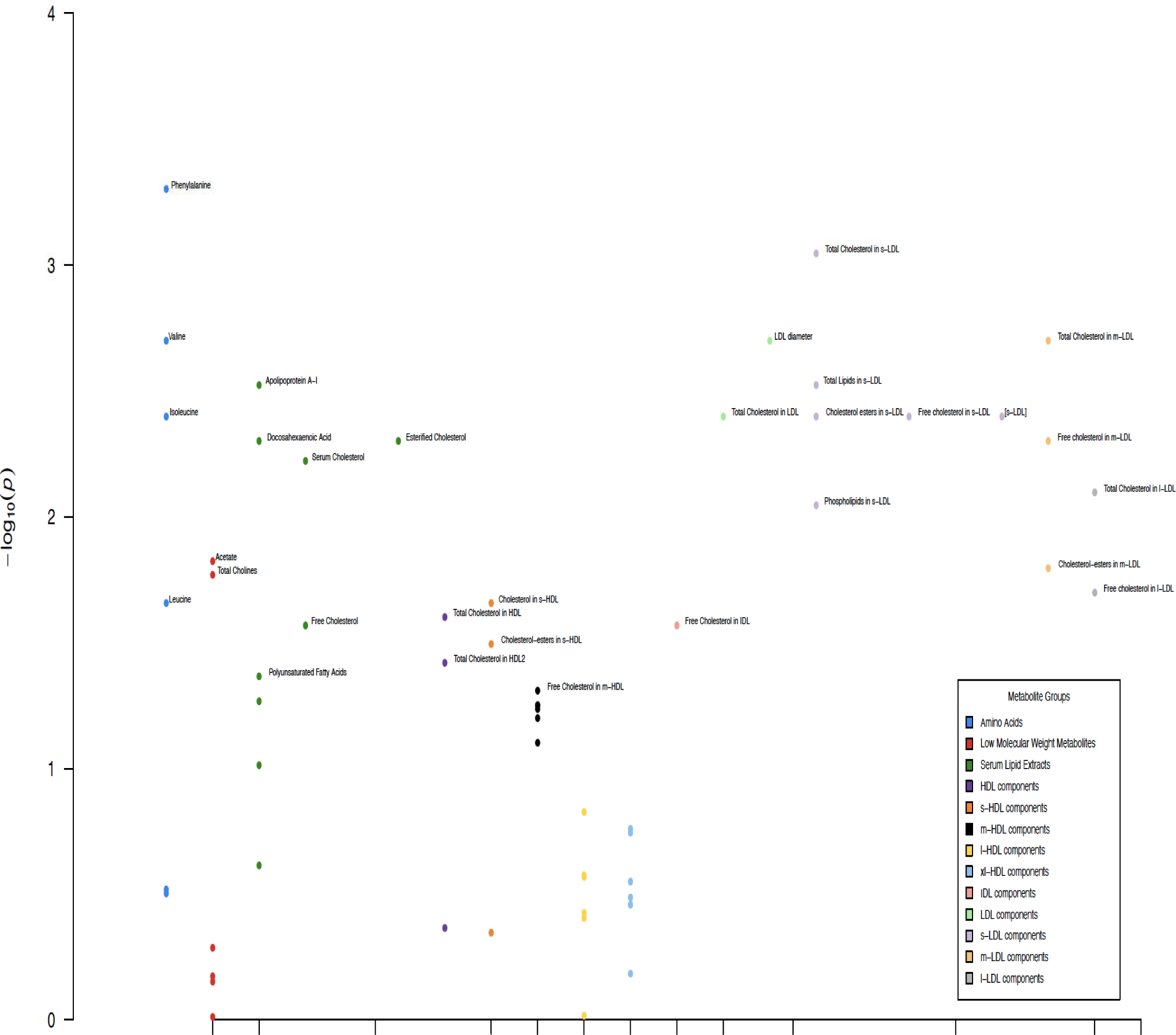
Manhattan Plot of associations between Metabolites and Snoring. Metabolites with a significant association with snoring in Model 2 are annotated HDL, high-density lipoprotein; s-HDL, small-HDL; m-HDL, medium-HDL; l-HDL, large-HDL, xl-HDL, extra-large HDL; IDL, intermediate-density lipoprotein; LDL, low-density lipoprotein; s-LDL, small-LDL; m-LDL, medium-LDL; l-LDL, large-LDL; CE, cholesterol-esters; FC, free cholesterol;

**Figure 2.**
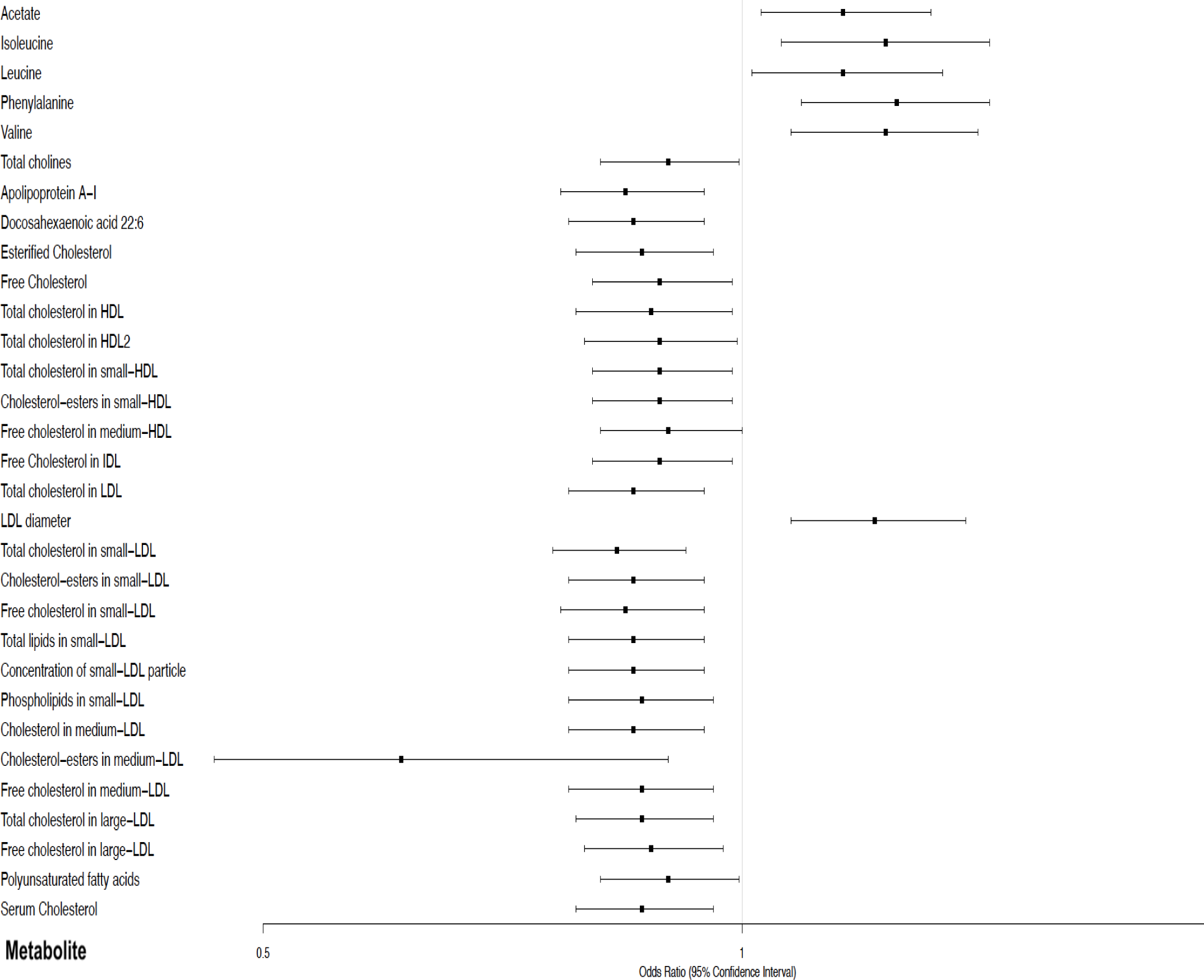
Forest plot of odds ratios and 95% confidence intervals of association for metabolites with snoring which were significant in Model 2 (adjusted for age, sex, ethnicity and years of education, waist-hip-ratio, cardiovascular disease, Type 2 Diabetes, hypertension medication, alcohol units and smoking status) OR, odds ratio; CI, confidence interval. Other abbreviations as in **Figure 1**.

### SENSITIVITY ANALYSIS

Additional metabolites which were associated in the fully-adjusted models but did not pass the screening stage were identified (**Supplementary Table S2**). A 1SD increase in glycine, free cholesterol and sphingomyelins was related to greater odds (ORs ≅ 1.15) of having difficulty falling asleep. Creatinine and valine were associated with lower odds (≅ 0.90) of early morning waking, while albumin and lactic acid associated with lower odds (≅ 0.87) of waking up tired. Free cholesterol, phospholipids and total lipids in medium and large LDL, small very low-density lipoprotein (VLDL) and very small VLDL; and apolipoprotein B and omega-3 fatty acids were associated with lower odds of snoring (≅ 0.90).

## 4. DISCUSSION

Using data from the SABRE cohort, we show in cross-sectional analysis, that 32 circulating plasma metabolites are associated with distinct sleep quality phenotypes. In particular, metabolites that were associated both with higher as well as lower odds of difficulty falling asleep and snoring were identified. Increased levels of some of the metabolites are associated with lower odds of early morning waking and waking up tired.

Difficulty falling asleep is one of the most frequent symptoms reported by insomnia patients^16^. Our results show that increased levels of histidine, isoleucine and valine were associated with lower odds of difficulty falling asleep even after adjusting for the relevant covariates. Interestingly, all three are essential amino acids, which means that they cannot be synthesized de novo and therefore, may be linked to a deficient diet. Histidine is a precursor of histamine which has been proposed as a regulator of wakefulness^17^. Isoleucine and valine are precursors of both glutamate and gamma-butyric acid^18^, the main excitatory and inhibitory neurotransmitters. By restoring the inhibition: excitatory ratios, their supplementation could restore normal sleep patterns. In this study, we report that BCCA are associated with lower odds (≅ 0.90) of difficulty falling asleep. To date, branched-chain amino acids such as leucine and valine have been successfully trialed to improve sleep quality in certain population groups such as cirrhosis patients^19^..

We also observed that in SABRE snoring was associated with 31 metabolites suggesting a potential complex metabolic disturbance. Most of these metabolites are from the lipid profile and include serum lipid extracts (such as DHA, polyunsaturated fatty acids) and cholesterol, cholesterol-esters and phospholipids in small/medium/large LDL and HDL fractions. Although the snoring-insulin resistance and associated dyslipidemia connection has previously been reported^7^, the novelty of this study comes from the detailed lipoprotein analysis which identified specific lipoprotein components. For example, the largest odds ratio for snoring was for cholesterol-esters in medium LDL. Our data show that both higher LDL and HDL associate with lower odds of snoring. The effect of HDL is consistent with the existing literature^7^, but the effect of LDL is not. NMR quantifies lipoproteins based on molecular weight and diameter into the lipoprotein subclasses. Given that HDL and LDL are so tightly correlated (mostly positively) (**Figure 3**), the negative association of LDL with snoring could be due to their tight relationship rather than a true effect. Although an inverse correlation between HDL and LDL is to be expected, it should be noted that lipoproteins in metabolism are in a constant state of flux with complex interactions, rather than discrete measures. Moreover, there is a shift from the traditional thinking that HDL is “good” and LDL is “bad” as there are beneficial subfractions of LDL and detrimental ones of HDL. In addition, the directionality of the associations between snoring, diabetes, WHR and metabolites is still a matter of debate as complex metabolic interactions exist (**Figure 4**).

**Figure 3.**
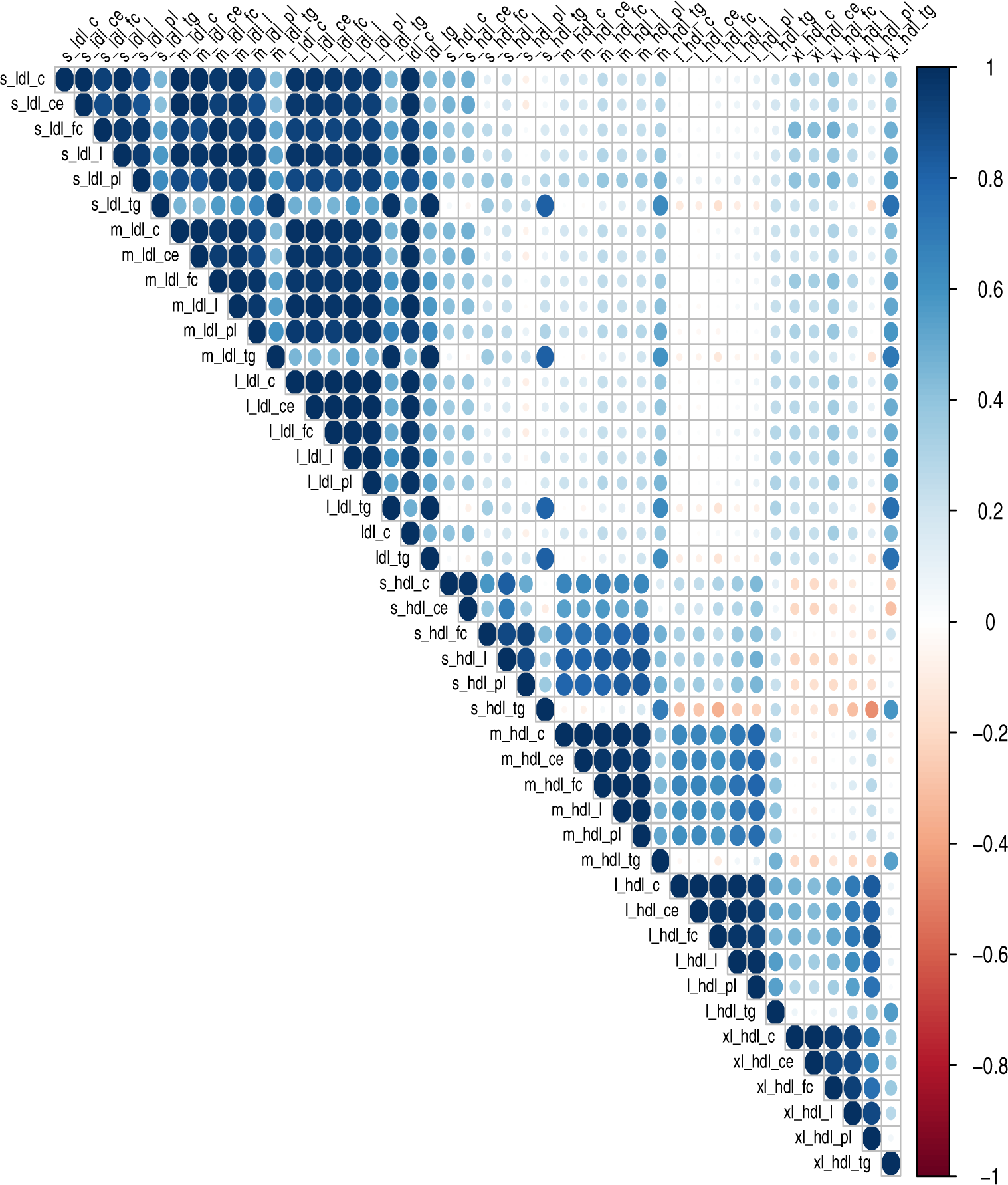
Correlation matrix of HDL and LDL sub-fractions Positive correlations are displayed in blue and negative correlations are displayed in red. Color intensity and the size of the circle is proportional to correlation coefficients. The legend color displays the correlation coefficients range and the corresponding colors. Abbreviations as in **Figure 1**.

**Figure 4.**
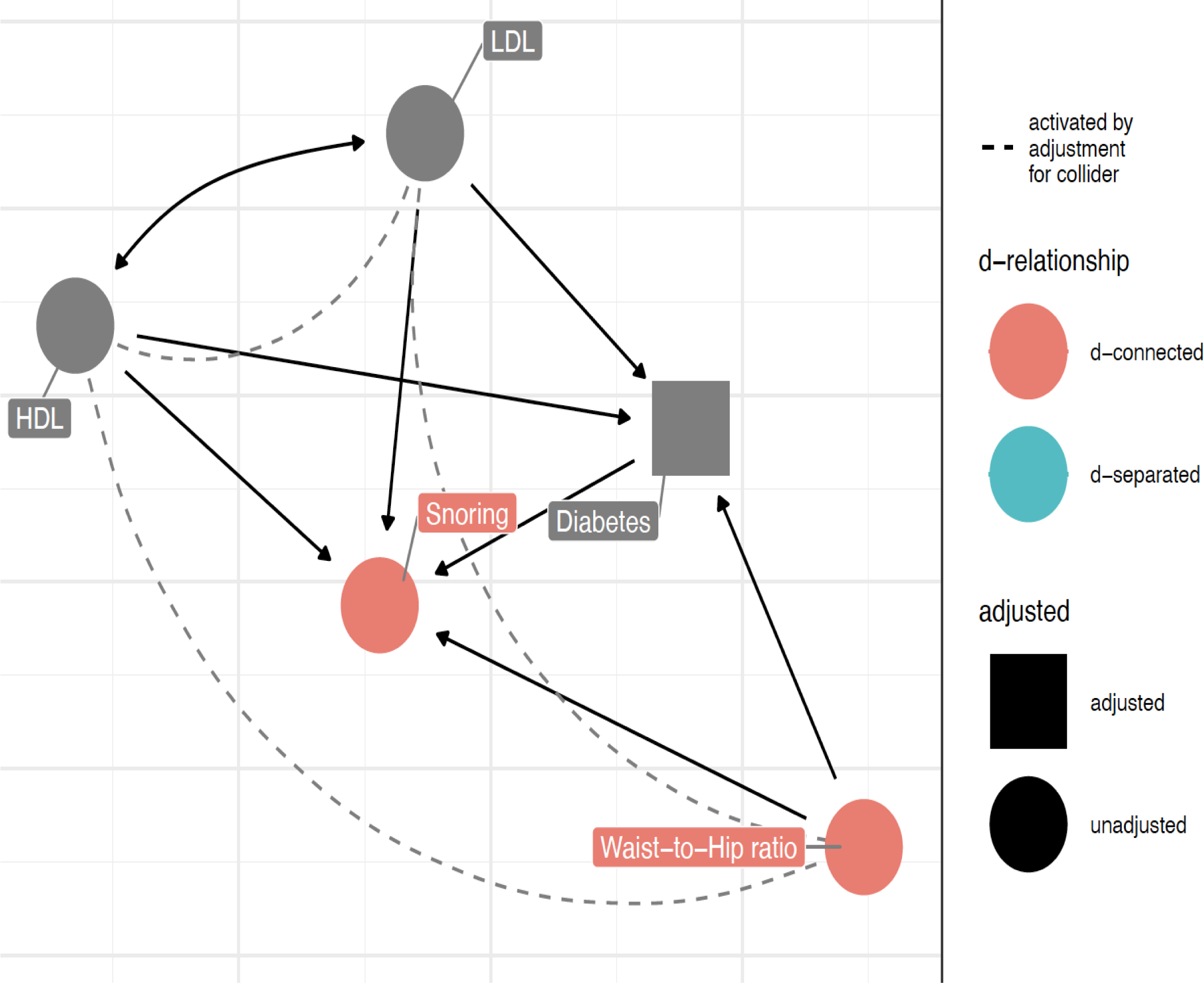
Directed Acyclic Graph for Snoring Low HDL, high LDL, high waist-to-hip (WHR) ratio and type 2 diabetes (T2DM) have been shown to be associated with snoring. However, a high WHR is associated with T2DM and both variables are associated with dyslipidemia. This direct acyclic graph highlights the complex interactions which exist within this system. Abbreviations as in **Figure 1**.

Lastly, the inverse association between DHA and snoring severity has previously been reported^20^. Beneficial effects of ω-3 fatty acids in terms of reduced daytime sleepiness have been observed after supplementation in deployed soldiers^21^. DHA is an important component of neural membranes which has been postulated to be both a synaptic and a neuromodulator altering the levels of glutamate, monoamines, acetylcholine and endocannabinoids^22^.

To the best of our knowledge, we are the first to report acetate, all BCCA and phenylalanine to be associated with snoring. As acetate generates acetyl-CoA, it has been proposed as an epigenetic metabolite to regulate lipid synthesis under hypoxia^23^ which can occur during snoring. Phenylalanine is a precursor to dopamine and norepinephrine which have been shown to be downregulated in sleep deprivation^24^. Interestingly, higher valine has been associated with snoring (higher odds), early morning waking (lower odds) and difficulty falling asleep (lower odds) making it more likely to be a key biological player in regulating sleep. Given the wide-ranging effects, it could be that valine operates within a narrow homeostatic window with higher levels promoting sleep (lower odds of difficulty falling asleep and early waking), but if they are too high snoring could arise. This theory is supported by its involvement in multiple important processes, such as protein synthesis, energy production, glutamate compartmentalization, and indirect control of serotonin, dopamine and noradrenaline neurotransmitter synthesis^25^.

Our sensitivity analysis revealed that higher creatinine and valine associate with lower odds of early morning waking. A positive relationship between creatinine and long sleep duration has been previously reported^26^. Creatinine is a breakdown product of creatine phosphate. The latter is of neurophysiological importance acting as an antioxidant, a neuromodulator (of Gamma aminobutyric acid A (GABA_A_) and of N-methyl-D-aspartate (NMDA) receptors) and a regulator of neuronal energy metabolism. It has been postulated to neutralize the negative effects of reactive oxygen species which occur on the background of chronic psychological stress^27^. To date, it has been successfully trialed to improve mood and performance following sleep deprivation^28^. Its potential for the prevention of early morning waking has not been yet explored in clinical studies.

Waking up tired has been associated with chronic fatigue syndrome. Our sensitivity analysis identified that higher albumin and lactic acid are related to lower odds of waking up tired. Lower albumin correlates with fatigue in chronic kidney disease patients^29^. We are the first to generalize this association in a large-scale, predominantly healthy cohort. Although we did not specifically measure tryptophan, the observed effect might be mediated by this albumin-bound amino acid, as it has been linked to a higher serotonin: dopamine ratio leading to central, as opposed to peripheral, fatigue ^30^. Serum lactate has previously been reported as a possible sleep/wake biomarker with higher levels during wakefulness; and persistent and sustained decline during nonrapid eye movement sleep^31^. Serum lactate could potentially be extended as a biomarker for the early morning waking phenotype. It could also be that central fatigue may stem from a lower neuronal glucose consumption translating into lower plasma lactate levels.

Strengths of the present study include the broad spectrum of metabolites enabled by NMR metabolomics, large sample size and extensive adjustment for covariates known to affect the relationship between metabolites and sleep. The sample is representative of primary care registration as healthcare services are free nationally in the UK. Limitations to the study include its inherent cross-sectional nature and the failure to capture longitudinal effects or to support causality. The directionality of the associations between metabolites and sleep phenotypes is still a matter of debate and whether metabolites cause or are a consequence of abnormal sleep has yet to be elucidated. In addition, there were only a few women included due to the study design. Another limitation is the long-term storage of samples (over 20 years) before NMR analysis.

## 5. CONCLUSION

Histidine, leucine and valine associated with lower odds of difficulty falling asleep, while branched chain amino acids were positively associated with snoring. Total cholesterol in certain HDL and LDL subfractions appeared beneficial in terms of snoring. Although the current evidence is unable to support causality, the identified metabolites could provide a direction for future studies to further understand abnormal sleep patterns.

## Data Availability

The data might be available upon reasonable request.

## Acknowledgements

The authors would like to thank all the SABRE members for their participation and continuous engagement with follow-up and all SABRE scientific and data collection teams.

